# Stroke Knowledge and Its Determinants Among At-Risk Middle-Aged Adults in a Semi-Urban Nigerian Community

**DOI:** 10.64898/2026.07.26.26358987

**Authors:** Ikeanyi Chinelo Nneka, Odeke Dorathy Akunna, Ugochukwu Ezinwanne Jane, Unyime Eshiet, Isah Abdulmuminu, Ubaka Chukwuemeka Michael

## Abstract

**Background:** Stroke is a leading cause of premature mortality and disability in sub-Saharan Africa. Effective prevention depends on timely recognition of stroke symptoms and awareness of its risk factors. This study assessed stroke knowledge and its independent predictors among at-risk middle-aged adults in a semi-urban Nigerian community.

**Methods:** A community-based cross-sectional study was conducted among 316 adults aged 40–65 years using interviewer-administered questionnaires. Stroke knowledge was defined as adequate recognition of both stroke symptoms and risk factors. Pearson’s chi-square test and multivariable binary logistic regression were used to identify independent predictors of combined stroke knowledge.

**Results:** Although 296 (93.7%) respondents were aware of stroke, only 112 (35.4%) demonstrated good combined knowledge of stroke symptoms and risk factors. Sudden weakness of face, arm, or leg was the most correctly identified symptom (78·6%), while sudden visual disturbance (34·1%) and severe headache (37·6%) were poorly recognized. Hypertension was the most identified risk factor (87·6%), but atrial fibrillation (32·9%), alcohol (51·4%), and diabetes (57·0%) were inadequately recognized. Good heart disease knowledge (aOR 3.09, 95% CI 1.82–5.28; p < 0.001) and a previous diagnosis of hypertension (aOR 2.32, 95% CI 1.14–4.71; p = 0.020) were independently associated with good combined stroke knowledge, whereas post-secondary education (aOR 0.37, 95% CI 0.15–0.93; p = 0.034) was inversely associated.

**Conclusion:** Despite high stroke awareness, comprehensive knowledge of stroke symptoms and risk factors remains suboptimal among at-risk middle-aged adults. Integrating stroke education into existing cardiovascular prevention services may provide a practical strategy for improving stroke literacy in high-risk populations.

---

**Keywords:** cardiovascular disease; community-based survey; health literacy; Nigeria; stroke; stroke awareness.

## Introduction

Stroke remains one of the leading causes of death and disability worldwide and is the third leading cause of death globally, accounting for approximately 7.3 million deaths and 11.9 million incident cases in 2021, with sub-Saharan Africa bearing one of the highest regional burdens where annual incidence rates are estimated to be as high as 316 per 100,000 population.[1,2] A systematic review and meta-analysis estimated the pooled crude prevalence of stroke in Nigeria at 6.7 per 1,000 population, with an annual incidence of approximately 26 per 100,000 person-years, The review also report that stroke also constitutes a major cause of neurological admissions and mortality in Nigerian hospitals, reflecting its substantial health-system burden.[3] A community-based study in Nigeria reported a much higher stroke prevalence of 1,460 per 100,000 population, among the highest documented globally, underscoring the growing burden of stroke and the need for effective prevention strategies.[4]

Up to 80% of strokes are considered preventable through control of modifiable vascular risk factors, including hypertension, diabetes mellitus, dyslipidemia, obesity, smoking, unhealthy diet, and physical inactivity.[1] Despite this, stroke outcomes in Nigeria, remain poor, with studies reporting that the majority of patients present late to hospital often beyond 24 hours of symptom onset and up to 85% presenting outside the early therapeutic window. This delay has been linked to low public awareness of stroke warning signs, poor health literacy, and non-hospital first contact patterns.[5]

Effective stroke prevention depends not only on controlling vascular risk factors but also on public recognition of stroke symptoms and timely care-seeking. Stroke management is critically time dependent and early recognition of stroke symptoms is critical for survival. Public knowledge of stroke warning signs captured by the B.E.F.A.S.T. acronym (Balance loss, Eyes/vision changes, Face drooping, Arm weakness, Speech difficulty, Time to call emergency services) is a direct determinant of emergency response time and treatment-seeking behaviour.[6,7] Similarly, knowledge of modifiable risk factors (hypertension, diabetes, dyslipidaemia, obesity, physical inactivity, smoking, and excessive alcohol use) is essential for primary prevention and lifestyle modifications.[8,9] Stroke literacy remains suboptimal across Nigeria and other parts of sub-Saharan Africa. In Nigeria, a study among adults at high risk for stroke found that only 39.6% could identify at least one stroke warning sign, while 60.4% could not identify any warning sign.[10] Similarly, a community survey in a southwestern university in Nigeria found that although many respondents recognized individual warning signs and risk factors, few demonstrated comprehensive knowledge of multiple stroke warning signs and modifiable risk factors.[11] Comparable findings have been reported in Ghana, where 73.1% of respondents had inadequate knowledge of stroke warning signs and similar percent had inadequate knowledge of stroke risk factors.[12] Comparable findings have also been reported in Ethiopia and Uganda, where stroke awareness and recognition of warning signs remain poor.[13,14]

Stroke and cardiovascular disease share many modifiable vascular risk factors and knowledge of heart disease may influence stroke literacy. Timely identification and effective control of these vascular risk factors are central to the primary and secondary prevention of strokes, highlighting the importance of public awareness and risk-factor modification in reducing stroke burden.[15] A systematic review and meta-analysis of combined cardiovascular and cerebrovascular educational-behavioral programs for culturally and linguistically diverse communities found that such multi-component and usually group-based improved CVD or stroke knowledge improved.[16] Therefore, heart disease awareness is an important covariate in analyses of stroke knowledge, and understanding its independent contribution alongside sociodemographic factors strengthens both scientific understanding and policy design.

Middle-aged adults represent an important target population in whom stroke primary prevention is most beneficial but also most neglected, as nearly 38% of strokes occur in this age group.[17] Most individuals in this age group are in their active years and a stroke event results in severe social and economic consequences, including increased disability and lost productivity.[18] Therefore, prevention of stroke is typically economically beneficial and results in reduced economic burden on households and society due to the reduction in the cost of productivity loss and cost of disability from the condition. Despite being an important target population, middle-aged adults living in semi-urban communities such as Nsukka may have limited access to specialist cardiovascular and neurological services. Combined with increasing cardiovascular risk factors associated with urbanization, this makes community-based stroke awareness particularly important.

Much of the existing Nigerian literature on stroke knowledge has focused on hospital-based patients [19,20] and healthcare workers [21–23] whereas community-based studies remain relatively few. Furthermore, evidence on stroke awareness among community-dwelling adults, particularly those at elevated risk of stroke in semi-urban settings, remains limited. The aim of this study was to assess stroke knowledge, defined as recognition of stroke symptoms and awareness of stroke risk factors, among at-risk middle-aged adults in a semi-urban Nigerian community. The secondary objective was to identify independent predictors of stroke knowledge, including knowledge of heart disease, to explore the interconnected nature of cerebrovascular and cardiovascular health literacy.

## Materials and Methods

### Study design and setting

The community-based cross-sectional study was conducted in Nsukka Local Government Area of Enugu State, southeastern Nigeria. According to the 2006 National Population Census, the LGA had a population of 309,448, with subsequent projections estimating a population of approximately 444,100 residents by 2022. The largely sub-urban center (around the University of Nigeria campus) and rural population comprises civil servants, traders, artisans, students, and informal sector workers. Nsukka exhibits many of the characteristics of a typical Nigerian semi-urban community undergoing epidemiological transition, with increasing prevalence of hypertension and other cardiovascular risk factors. As in many semi-urban settings in Nigeria, access to specialized cardiovascular and neurological services is concentrated in larger urban centers, highlighting the importance of community-based prevention and health literacy interventions.

### Population and sampling

The study population for this study comprised adults aged between 40 and 65 years (identified as “middle-aged” for this study) who were permanent residents of the community. These participants should have resided in the community for at least twelve months and provided informed consent before being included in this study. Individuals with severe cognitive impairment, acute illness or blatantly refused to participate at the time of interview were excluded.

The sample size of participants for the study was determined using the Leslie–Kish formula for estimating a single population proportion in a cross-sectional survey:

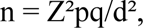

where Z = 1.96 (95% confidence level), p = 0.30 (based on previous evidence of adequate stroke knowledge among Nigerian adults), q = 1 − p = 0.70, and d = 0.05 (margin of error). This yielded a minimum sample size of 323 participants. To account for an anticipated non-response rate of 7%, the sample size was adjusted upward to 347 participants.

Accordingly, 347 questionnaires were distributed to eligible participants. Following data collection and quality assurance procedures, 316 questionnaires (representing a completion rate of 91.1%) were retained for analysis after excluding incomplete responses and records with substantial missing data.

A multistage sampling technique was employed to recruit study participants. In the first stage, enumeration areas and streets within Nsukka town were identified using records from the Nsukka Local Government Town Planning Department. Selected streets were chosen by simple random sampling. In the second stage, households within each selected street were sampled using proportionate systematic random sampling. Finally, one eligible adult was recruited from each sampled household. Where multiple eligible adults resided in the same household, the household head was selected; in the absence of the household head, the most senior eligible adult available at the time of the survey was recruited.

### Data collection instrument

Study data were collected using a structured interviewer-administered questionnaire adapted from previous cardiovascular and stroke awareness surveys. The instrument comprised four sections. Section A obtained respondents’ sociodemographic characteristics. Section B assessed the respondents’ awareness of stroke (i.e. if respondents had heard of stroke or knew anyone who had stroke) and knowledge of heart disease which was assessed using an abbreviated version of the validated 30-item Heart Disease Fact Questionnaire (HDFQ).[24] Sixteen items were purposively selected to retain representation of the major knowledge domains of the original instrument, including cardiovascular risk factors, preventive behaviors, heart attack symptoms, cardiovascular physiology, and common misconceptions. Section C evaluated respondents’ knowledge of eight stroke symptoms (of which three are distractor symptoms); with responses to each item classified either as “a stroke symptom”, “not a symptom”, or “unknown” (CDC 2008).^25^ Section D examined respondents’ knowledge of thirteen stroke risk factors (of which one is a distractor risk factor) with the item statements rated as either “a risk factor”, “not a risk factor”, or “unknown”.[8]

The questionnaire was pilot-tested among 20 community-resident university staff who were not included in the main study to assess the clarity, comprehensibility, and appropriateness of the items. Minor revisions were made based on participant feedback, including the addition of the phrase “irregular heartbeat” alongside “atrial fibrillation” to improve comprehension of this stroke risk factor among lay respondents.

### Outcome variable

The primary outcome was “combined stroke knowledge”. Participants were classified as having good, combined stroke knowledge if they correctly identified at least 70% of the listed stroke symptoms and at least 70% of the listed stroke risk factors, corresponding to a minimum of 6 out of 8 symptom items and 9 out of 13 risk factor items, respectively. The 70% threshold was chosen a priori to reflect a satisfactory level of knowledge across both domains and to distinguish participants with consistently high awareness from those with partial knowledge thus reflecting the dual competencies required for effective primary prevention and emergency recognition.

### Predictor variables

Independent variables included: sex, age group, highest educational attainment, marital status, employment status, monthly income, self-reported medical diagnosis and heart disease knowledge (good [≥70% i.e. 11/16 items] vs. poor).

### Statistical analysis

Completed questionnaires were reviewed for completeness, consistency, and eligibility prior to analysis. A total of 316 questionnaires met the inclusion criteria and were coded and entered into IBM SPSS Statistics version 27.0 (IBM Corp., Armonk, NY, USA) for analysis.

Descriptive statistics were used to summarize participants’ sociodemographic and clinical characteristics. Categorical variables were presented as frequencies and percentages.

The primary outcome was combined stroke knowledge, defined as correct identification of at least 70% of both the stroke symptom items (≥6 of 8 items) and the stroke risk factor items (≥9 of 13 items). Participants meeting both criteria were classified as having good combined stroke knowledge, while all others were classified as having poor combined stroke knowledge. Heart disease knowledge was evaluated as an explanatory variable and categorized as good or poor according to the predefined scoring criteria.

Associations between explanatory variables and combined stroke knowledge were initially assessed using Pearson’s chi-square test. Variables demonstrating statistical significance at the bivariate level (p < 0.05) were subsequently entered into a multivariable binary logistic regression model to identify independent predictors of combined stroke knowledge. Results are presented as adjusted odds ratios (aORs) with corresponding 95% confidence intervals (CIs). All statistical tests were two-tailed, and statistical significance was set at p < 0.05.

### Ethical considerations

Ethical approval was obtained from the Faculty of Pharmaceutical Sciences Ethics Committee, University of Nigeria (UNNFP-HREC) prior to data collection. Community and household-level entry permissions were obtained from the local government authorities. All participants provided written verbal consent during each visit. Participation was voluntary, and confidentiality and anonymity of respondents were maintained throughout the study. The study was conducted in accordance with the ethical principles of the Declaration of Helsinki (2013 revision).

## RESULTS

### Sociodemographic and clinical characteristics

A total of 316 respondents were included in the final analysis. Females constituted 51.3% (n = 162) of the sample. The most represented age group was 40–45 years (n = 124; 39.2%), and the majority (n = 290; 91.8%) had attained post-secondary education, reflecting the university town context of Nsukka. Most respondents were married (n = 192; 60.8%) and employed (n = 223; 70.6%). Monthly income was broadly distributed, with 19.9% earning below ₦50,000, 23.1% in the ₦50,000–100,000 range, 27.2% in the ₦100,000–150,000 range, and 24.1% earning above ₦150,000. Self-reported hypertension was the most common cardiovascular diagnosis (n = 55; 17.4%), followed by diabetes (n = 11; 3.5%) and hypercholesterolemia (n = 8; 2.5%). The majority (n = 219; 69.3%) reported no cardiovascular diagnosis. Most respondents had heard of stroke (n = 296; 93.7%), and 82.9% (n = 262) knew someone personally affected by stroke. Full sociodemographic characteristics are presented in **Table 1**.

**Table 1.**
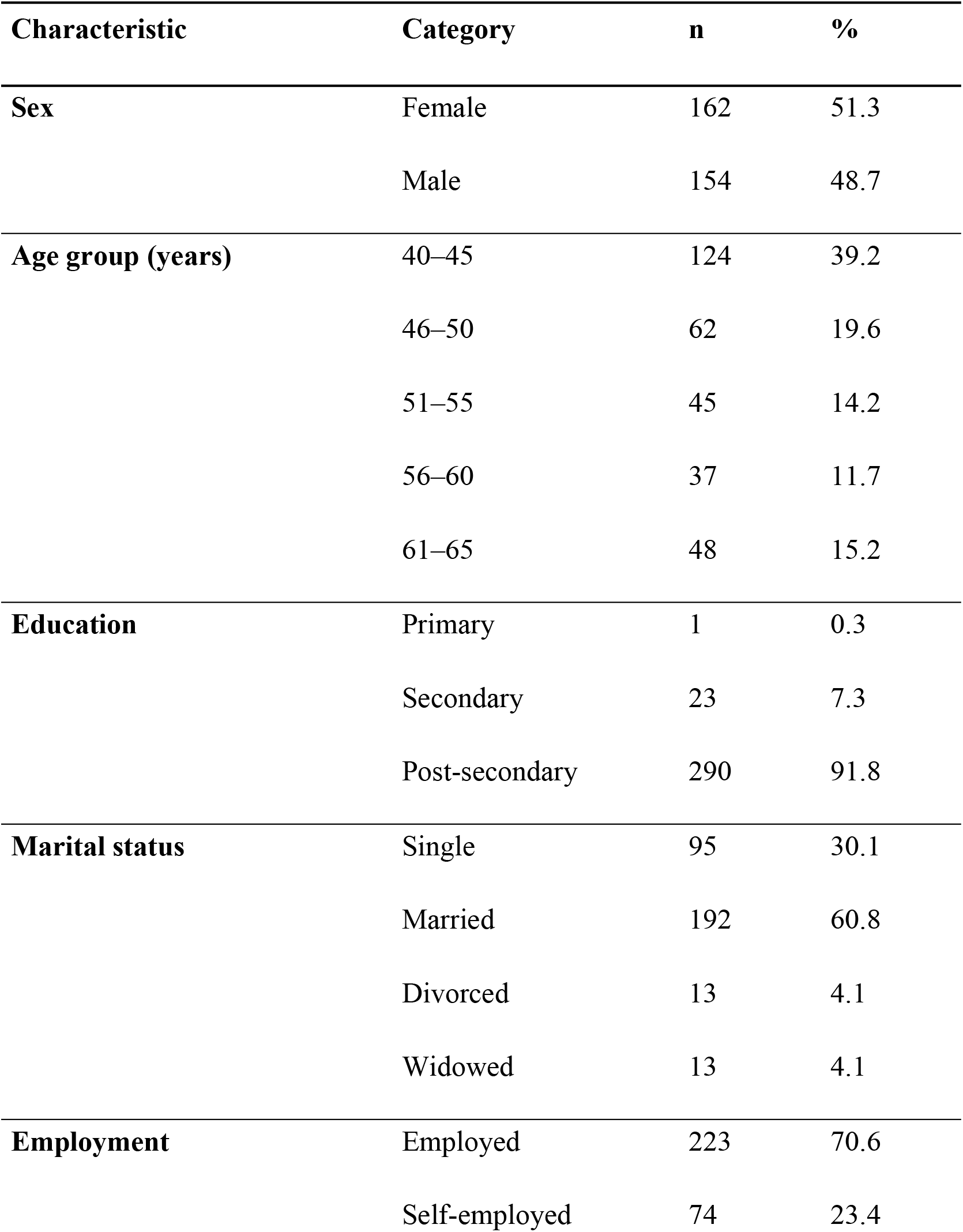
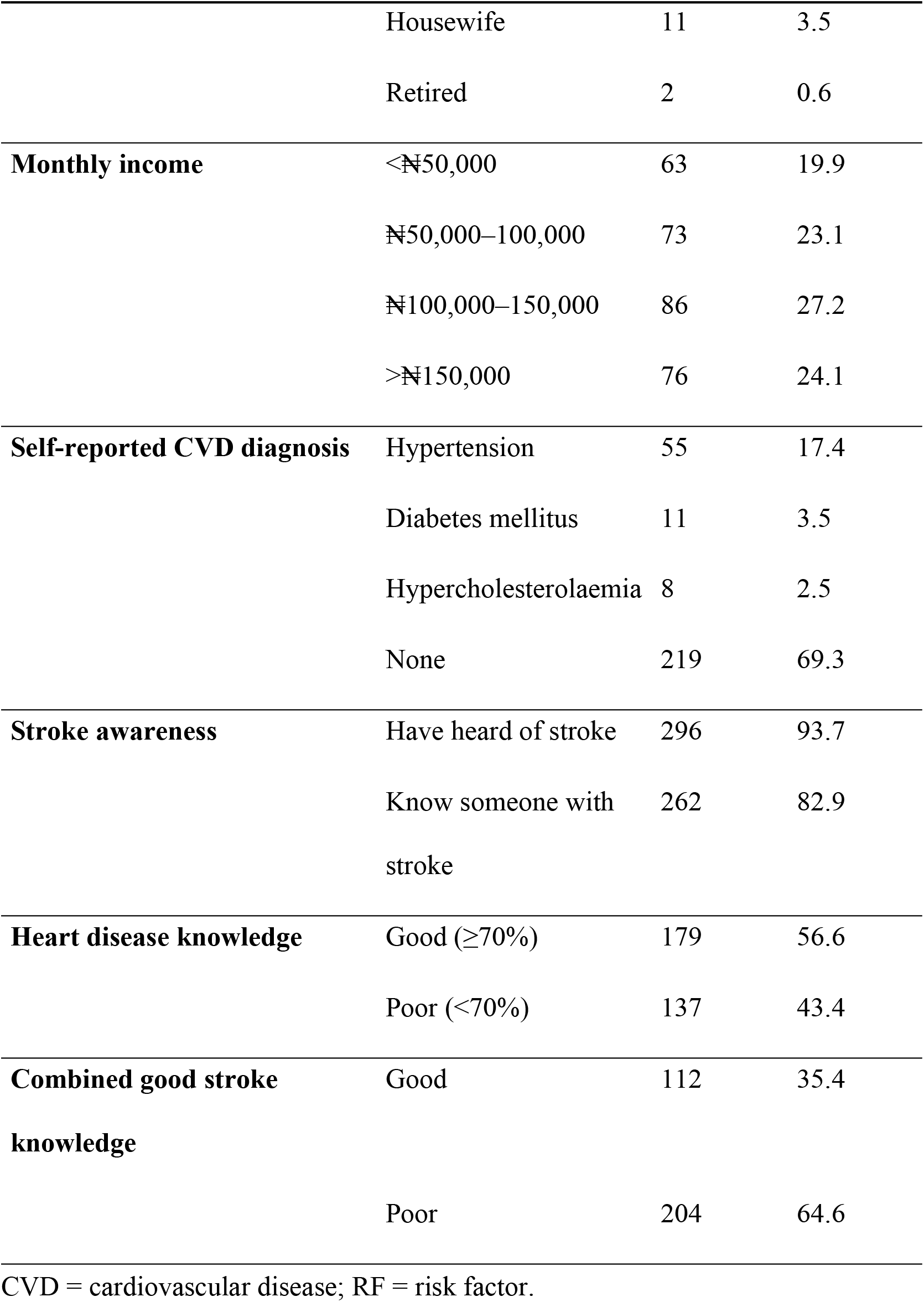
Sociodemographic and clinical characteristics of respondents (N=316)

### Knowledge of stroke symptoms

The correct identification of specific stroke symptoms was variable and suboptimal (**Table 2**). The most recognized genuine stroke symptom was sudden numbness or weakness of the face, arm, or leg (n = 246; 78.6%), followed by sudden trouble walking, dizziness, or loss of balance (n = 213; 68.1%) and sudden confusion or trouble speaking (n = 199; 63.2%). By contrast, two other genuine stroke symptoms i.e. sudden visual disturbance in one or both eyes (n = 107; 34.1%) and sudden severe headache with no known cause (n = 117; 37.6%), were recognized by fewer than two in every five respondents. Notably, the three non-stroke (distractor) symptoms were incorrectly endorsed by substantial proportions: sudden nosebleed (52.2%), high temperature (49.8%), and sudden vomiting (5.8%). The mean stroke symptom knowledge score was 3.86 ± 1.65 out of 8 correctly classified items. When we applied the ≥70% threshold to stroke symptom knowledge alone, 182 respondents (57.6%) demonstrated adequate symptom recognition.

**Table 2.** Knowledge of stroke symptoms among middle-aged respondents in semi-urban settings (N=316)

| Common symptom? | Correctly<br>classified n<br>(%) | Incorrectly<br>classified n<br>(%) |
| --- | --- | --- |
| **Genuine stroke symptoms** |  |  |
| Sudden confusion / trouble speaking or understanding speech (Yes) | 199 (63.2) | 117 (37.0) |
| Sudden numbness or weakness of face, arm, or leg (Yes) | 246 (78.6) | 70 (22.1) |
| Sudden trouble seeing in one or both eyes (Yes) | 107 (34.1) | 209 (66.2) |
| Sudden trouble walking, dizziness, or loss of<br>balance/coordination (Yes) | 213 (68.1) | 103 (32.7) |
| Sudden severe headache with no known cause (Yes) | 117 (37.6) | 199 (63.0) |
| **Distractor symptoms** |  |  |
| Sudden nosebleed (No) | 163 (52.2)<sup>†</sup> | 153 (48.4) |
| Sudden vomiting (No) | 18 (5.8)<sup>†</sup> | 298 (94.3) |
| High temperature / fever (No) | 157 (49.8)<sup>†</sup> | 159 (50.2) |
| Mean symptom score (out of 8 correct classifications) | 3.86 ± 1.65 | — |

<sup>†</sup> For these items, "correctly classified" means respondents correctly identified that the item is NOT a stroke symptom.

### Knowledge of stroke risk factors

Correct identification rates for stroke risk factors are detailed in **Table 3**. Hypertension was the most consistently recognized risk factor (n = 275; 87.6%), followed by heart disease (n = 224; 71.3%) and stress (n = 220; 70.3%). However, recognition of other established risk factors was considerably lower: “irregular heartbeat” (atrial fibrillation) was identified by only 104 respondents (32.9%), alcohol by 161 (51.4%), and diabetes by 179 (57.0%). Unhealthy diet (52.2%) and smoking (58.0%) were also poorly recognized as independent stroke risk factors. A distractor item, cough as a risk factor, was incorrectly endorsed by 207 respondents (65.9%), indicating poor discriminative knowledge. The mean risk factor knowledge score was 7.98 ± 3.17 out of 13. Using the ≥70% threshold, 160 respondents (50.6%) demonstrated adequate risk factor knowledge. The proportion of achieving combined adequate knowledge of both stroke symptoms and risk factors was 112 (35.4%).

**Table 3.**
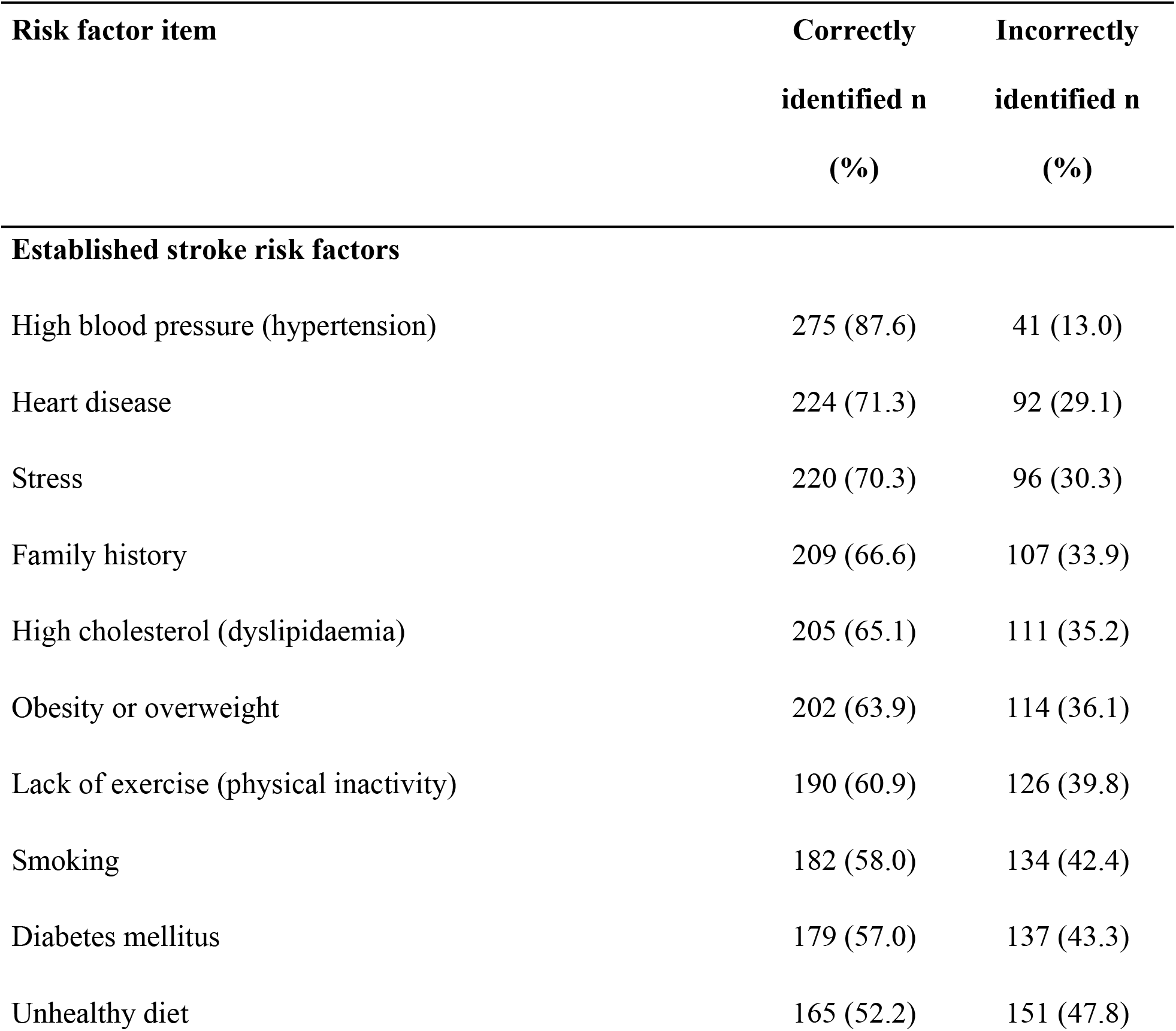
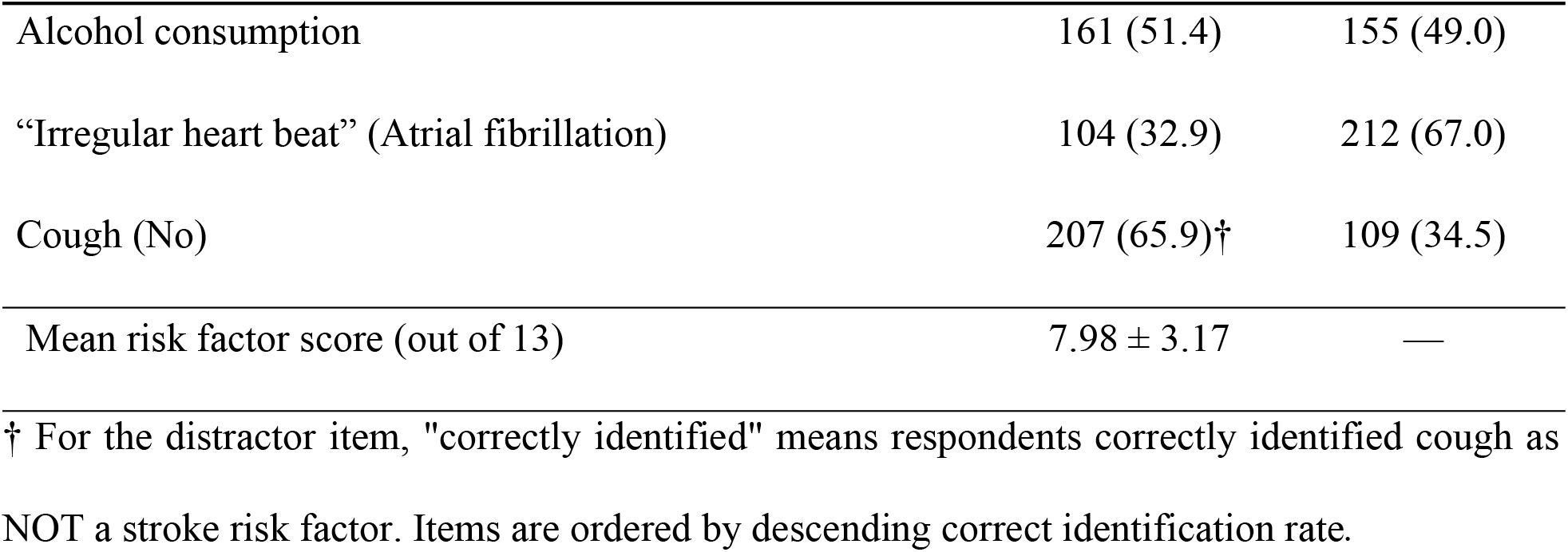
Knowledge of stroke risk factors among middle-aged respondents in semi-urban settings (N=316)

### Factors associated with good stroke knowledge

Bivariate associations between participant characteristics and combined good stroke knowledge are presented in **Table 4**. Sex was significantly associated with combined stroke knowledge, i.e., male respondents were more likely to have good knowledge than females (42.2% vs. 29.0%; χ² = 5.45, p = 0.020). Good heart disease knowledge was the most strongly associated factor (good knowledge group: 45.3% vs. poor knowledge group: 22.6%; χ² = 16.39, p < 0.001) for stroke knowledge. Good stroke knowledge was significantly more prevalent among participants with a prior hypertension diagnosis compared with those without hypertension (50.9% vs. 32.2%; χ² = 6.17, p = 0.013). Educational attainment showed a significant inverse association with combined stroke knowledge, with respondents having secondary or primary education demonstrating a higher prevalence of good knowledge than those with post-secondary education (65.2% vs. 33.1%; χ² = 11.39, p = 0.003). Monthly income was marginally significant (χ² = 8.08, p = 0.044), with lower income groups showing higher rates of good knowledge. Respondents’ age group, marital status, and employment were not significantly associated with stroke knowledge.

**Table 4.**
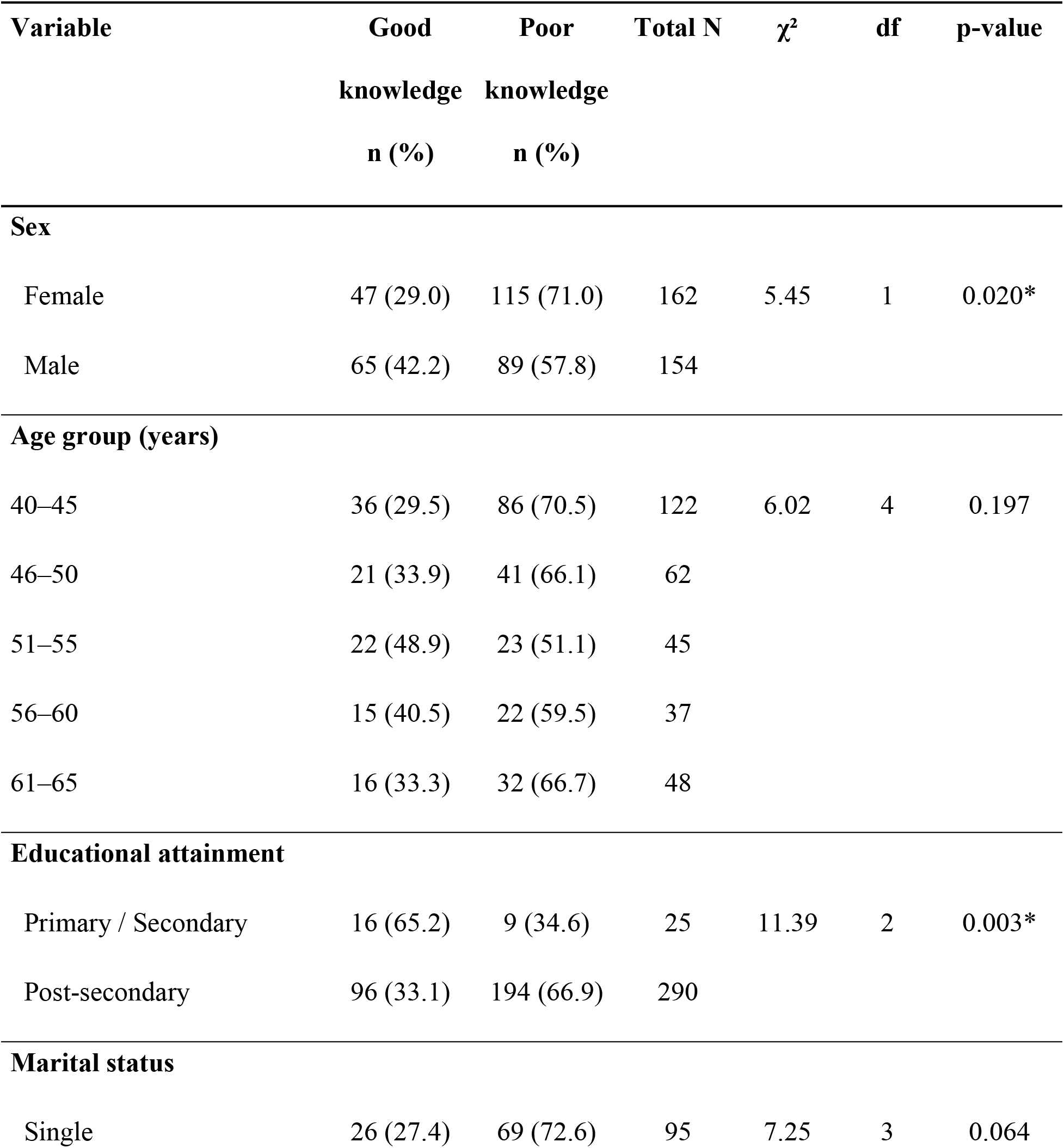
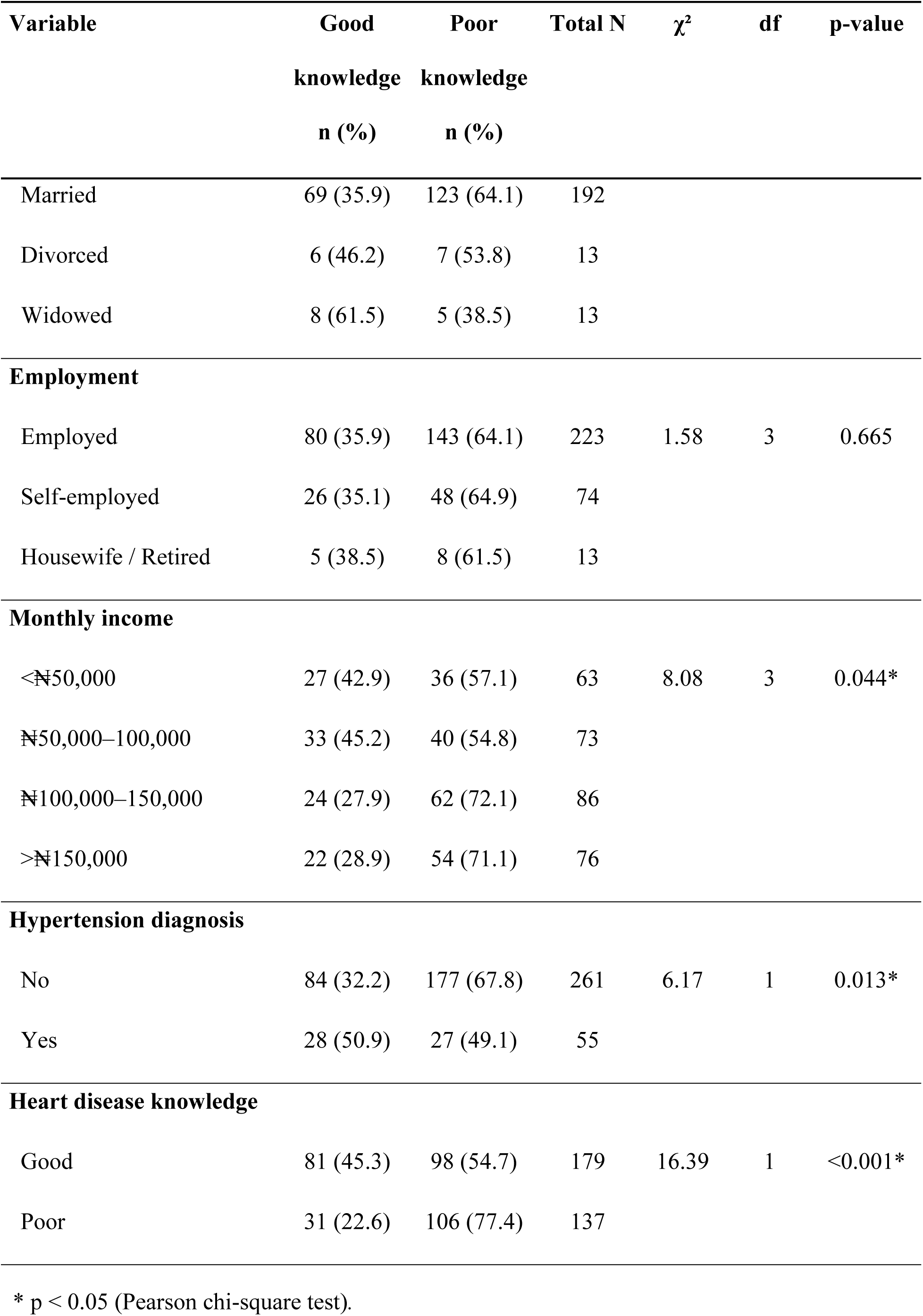
Bivariate analysis of factors associated with combined good stroke knowledge (N=316)

### Predictors of good stroke knowledge

**Table 5** presents the findings of the multivariable logistic regression analysis. After adjusting for all covariates, three variables independently predicted combined good stroke knowledge. Good heart disease knowledge was the strongest predictor (aOR 3.09, 95% CI 1.82–5.28; p < 0.001) as respondents with adequate heart disease knowledge were more than three times as likely to demonstrate good stroke knowledge. Prior hypertension diagnosis independently more than doubled the odds of good stroke knowledge (aOR 2.32, 95% CI 1.14–4.71; p = 0.020). Respondents with post-secondary education had lower odds of attaining good combined stroke knowledge than those with primary or secondary education (aOR = 0.37, 95% CI: 0.15–0.93; p = 0.034). Sex, age group and income were not independently significant after adjustment. The overall model was significant (likelihood ratio χ² = 73.12, df = 11, p < 0.001) with moderate explanatory power (Nagelkerke R² = 0.28), indicating improved fit over the null model.

**Table 5.** Multivariable logistic regression identifying independent predictors of combined good stroke knowledge (N=316)

| Variable | aOR | 95% CI | p-value |
| --- | --- | --- | --- |
| **Sex** |  |  |  |
| **(ref: female)** |  |  |  |
| Male | 1.66 | 1.00–2.75 | 0.052 |
| **Age group** |  |  |  |
| **(ref: 40–45 years)** |  |  |  |
| 46–50 | 0.92 | 0.45–1.88 | 0.823 |
| 51–55 | 1.99 | 0.91–4.38 | 0.086 |
| 56–60 | 1.34 | 0.58–3.12 | 0.495 |
| 61–65 | 0.94 | 0.41–2.19 | 0.892 |
| **Educational attainment** |  |  |  |
| **(ref: secondary/primary)** |  |  |  |
| Post-secondary | 0.37 | 0.15–0.93 | 0.034* |
| **Monthly income** |  |  |  |
| **(ref: <N50,000)** |  |  |  |
| N50,000–100,000 | 0.85 | 0.42–1.72 | 0.660 |
| N100,000–150,000 | 0.60 | 0.29–1.21 | 0.154 |
| >N150,000 | 0.51 | 0.24–1.08 | 0.078 |
| **Hypertension diagnosis** |  |  |  |
| **(ref: no)** |  |  |  |
| Yes | 2.32 | 1.14–4.71 | 0.020* |
| **Heart disease knowledge** |  |  |  |
| **(ref: poor)** |  |  |  |
| Good | 3.09 | 1.82–5.28 | <0.001* |

aOR = adjusted odds ratio; CI = confidence interval; ns = not significant. Model includes all variables simultaneously. \* $p < 0.05$

## DISCUSSION

### Principal findings

This study demonstrated a generally high level of stroke awareness within the community; however, fewer than half of respondents had good knowledge of both stroke symptoms and risk factors. Knowledge was characterized by selective recognition of stroke features, with obvious focal neurological deficits and hypertension as a risk factor most frequently identified. Other symptoms such as visual disturbance and sudden severe headache, as well as risk factors such as atrial fibrillation, were substantially underrecognized.

Adequate knowledge of heart disease, prior hypertension diagnosis, and educational level were independently associated with good stroke knowledge. Notably, post-secondary education showed an inverse association with combined good stroke knowledge.

### Previous studies

The suboptimal awareness of stroke warning signs observed in this study is consistent with previous evidence from Nigeria and other low-and middle-income countries. In a community-based study conducted in Benin City, Nigeria, fewer than 40% of adults were able to identify at least one stroke warning sign, while only a small proportion could correctly identify three or more warning signs, indicating substantial deficiencies in stroke symptom recognition.[10] Similar findings have also been reported elsewhere. In a recent community-based survey conducted across the seven northeastern states of India, only 19.2% of participants were aware of the warning signs and symptoms of stroke.[26] Collectively, these findings indicate that although respondents were familiar with some of the classical manifestations of stroke, important gaps remain in their ability to accurately recognize the full spectrum of stroke symptoms. This suggests that public education should move beyond promoting general awareness to improving recognition of the diverse clinical presentations of stroke.

Recognition of sudden weakness and speech difficulty as the most frequently identified stroke symptoms is consistent with previous studies conducted in Ghana and Nigeria, where unilateral weakness/paralysis and speech impairment were among the most commonly recognized warning signs of stroke.[11,12] In contrast, visual disturbance and sudden severe headache were relatively poorly recognized in our study, suggesting that public awareness remains largely centered on the classic FAST symptoms, while less typical manifestations are frequently overlooked. This finding has important clinical implications because posterior circulation strokes commonly present with visual symptoms, dizziness or headaches and are more likely to be missed or recognized late than anterior circulation strokes, contributing to delays in diagnosis and treatment.[27,28]

Another notable finding was the incorrect attribution of distractor symptoms, including fever, sudden vomiting and nosebleeds, as manifestations of stroke by a substantial proportion of respondents. This suggests difficulty distinguishing true stroke symptoms from unrelated clinical conditions and highlights important gaps in stroke-specific health literacy. Taken together, these findings indicate that although respondents were familiar with some classical manifestations of stroke, important deficiencies remain in their ability to accurately recognize the full spectrum of stroke symptoms. Improving public understanding of the characteristic features of stroke is essential because accurate symptom recognition is a major determinant of timely healthcare seeking and early access to acute stroke treatment.

Awareness of stroke risk factors was likewise suboptimal in this study. Previous Nigerian studies have similarly reported incomplete recognition of major stroke risk factors among adults, indicating that important gaps in stroke health literacy persist despite general familiarity with the disease.[11,29] The predominance of hypertension as the most frequently recognized risk factor is also consistent with findings from community-based studies in Ghana and Nigeria, where hypertension was more readily identified than less visible or less publicized risk factors such as obesity, alcohol use and cardiac conditions.[12,29] Similar patterns have been reported in India, where hypertension was consistently the most commonly identified stroke risk factor among community respondents.[30,31] Collectively, these findings suggest that public education continues to emphasize hypertension while other clinically important but less visible risk factors receive comparatively little attention.

The poor recognition of atrial fibrillation (“irregular heartbeat”) as a stroke risk factor highlights an important gap in stroke-specific health literacy, given that atrial fibrillation is a major and potentially preventable cause of ischemic stroke.[32] Evidence from community-based screening studies demonstrates that opportunistic screening in primary care and community pharmacy settings can improve the detection of previously undiagnosed atrial fibrillation.[33,34] Based on this evidence, current European Society of Cardiology guidelines recommend opportunistic atrial fibrillation screening among adults aged ≥65 years and other high-risk populations to facilitate early diagnosis and stroke prevention.[35] Given the growing burden of cardiovascular disease in sub-Saharan Africa, improving public awareness of atrial fibrillation should complement opportunistic screening strategies. Integrating simple pulse assessment and atrial fibrillation education into primary healthcare and community pharmacy services may strengthen stroke prevention by facilitating earlier identification and referral of high-risk individuals.

Overall, our findings are consistent with reports from Nigeria and Ethiopia showing that adults in both community and clinical settings often demonstrate general awareness of stroke but inadequate knowledge of its specific warning symptoms and risk factors.[29,36] These findings indicate that future public health interventions should shift from simply increasing awareness of stroke towards improving actionable knowledge that enables individuals and their families to recognize stroke promptly and seek emergency care. This has important clinical implications because failure by patients or their family members to recognize stroke warning signs is a major contributor to prehospital delays, reducing the likelihood of timely hospital presentation and access to time-sensitive interventions that improve clinical outcomes. [30,37]

Adequate heart disease knowledge was independently associated with good combined stroke knowledge, highlighting the close relationship between cardiovascular and cerebrovascular health literacy. Because stroke and heart disease share many modifiable risk factors, including hypertension, diabetes, obesity and atrial fibrillation, greater knowledge of cardiovascular disease is likely to enhance recognition of stroke symptoms and risk factors. This finding is consistent with emerging evidence demonstrating that broader cardiovascular health literacy is associated with improved stroke knowledge and preventive behaviors.[38]^38^ Rather than addressing stroke in isolation, these findings support integrated cardiovascular education strategies that simultaneously promote awareness of heart disease and stroke, particularly in resource-constrained settings where coordinated health promotion may be more efficient and sustainable.

Participants with a previous diagnosis of hypertension were also more likely to demonstrate good combined stroke knowledge. This finding is consistent with evidence that individuals living with hypertension have greater exposure to cardiovascular risk counselling during routine clinical care and therefore tend to possess better knowledge of stroke and its prevention than individuals without diagnosed hypertension, although important knowledge gaps often remain.[39] Routine hypertension care represents an important opportunity to integrate structured stroke education into chronic disease management. In Nigeria, regular interactions with primary healthcare providers and community pharmacists provide practical opportunities to reinforce stroke prevention messages and improve symptom recognition.

An unexpected finding was the inverse association between post-secondary education and good combined stroke knowledge. This contrasts with the broader literature, which consistently identifies higher educational attainment as a positive predictor of stroke knowledge. [36,40] The finding should therefore be interpreted cautiously. Respondents with post-secondary education constituted more than 90% of the study population, resulting in limited variability in educational attainment and a comparatively small reference group. In addition, the stringent composite definition of good stroke knowledge used in this study differed from that employed in many previous studies. These methodological considerations may partly explain the observed association, which should be confirmed in more educationally diverse populations.

#### Policy implications

The findings of this study have several implications for public health policy and practice. First, while general stroke awareness is relatively high, the persistence of gaps in specific symptoms and risk factor knowledge suggests that current public health messaging may be too narrow in scope. There is a need to strengthen educational interventions that move beyond general awareness to include comprehensive stroke symptom recognition, particularly less common but clinically important presentations.

Second, the strong association between hypertension diagnosis and stroke knowledge highlights the importance of integrating stroke education into routine hypertension care. This represents a practical opportunity to leverage existing health service contacts to improve stroke literacy among high-risk individuals.

Third, the low recognition of atrial fibrillation as a risk factor suggests the need for targeted educational strategies addressing non-hypertensive causes of stroke. This should be complemented by strengthening opportunistic screening approaches in primary care and community pharmacy settings, where feasible, to improve early detection.

Fourth, the observed association between heart disease knowledge and stroke knowledge supports the case for integrated cardiovascular education programmes rather than disease-specific messaging. Such an approach may be more efficient and sustainable within resource-limited health systems.

Finally, the findings support leveraging existing primary healthcare platforms, including community pharmacies, to deliver structured cardiovascular and stroke education. In many Nigerian communities, community pharmacists are readily accessible and frequently interact with individuals with hypertension and other cardiovascular risk factors. Incorporating brief, standardized counselling tools into routine practice may therefore represent a cost-effective strategy for improving stroke literacy and promoting early recognition and referral.

### Strengths and limitations

A major strength of this study is the comprehensive assessment of stroke literacy through simultaneous evaluation of both stroke symptom recognition and risk factor awareness using a composite outcome. Focusing on community-dwelling middle-aged adults at increased cardiovascular risk also provides evidence from an underrepresented population and enables identification of clinically relevant predictors, including cardiovascular disease knowledge.

This study has several limitations. Its cross-sectional design precludes causal inference. The predominance of respondents with post-secondary education may limit the generalizability of the findings to populations with more diverse educational profiles and may have influenced the observed association between educational attainment and stroke knowledge. In addition, knowledge and medical history were self-reported and therefore subject to recall and social desirability bias.

## CONCLUSION

This study demonstrates that although general awareness of stroke is high among at-risk middle-aged adults in this semi-urban Nigerian community, comprehensive knowledge of stroke symptoms and risk factors remains suboptimal. Adequate heart disease knowledge and a previous diagnosis of hypertension were independently associated with better combined stroke knowledge, highlighting the close relationship between cardiovascular and cerebrovascular health literacy. Improving stroke literacy may be achieved more effectively by leveraging existing cardiovascular prevention services than by relying solely on population-wide awareness campaigns. Integrating stroke education into routine cardiovascular care and primary healthcare services offers a practical and sustainable strategy for improving stroke literacy among individuals at greatest risk. Further studies are needed to validate the observed association between educational attainment and stroke knowledge in more diverse populations.

## Acknowledgement

The authors especially thank the Nsukka Local Government Council for the permission to carry out the survey.

## Data Availability

The anonymized dataset used in this study is available from the corresponding author upon reasonable request.

## Ethical Approval

Ethical approval was obtained from the University of Nigeria Teaching Hospital Health Research Ethics Committee (UNTH-HREC/2025/012). All participants provided written informed consent. The study was conducted in accordance with the Declaration of Helsinki (2013 revision).

## Author Contributions

Study conception and design: ODA, UEJ, UE, CMU. Data collection: ODA, IC, UEJ. Data analysis: IA, CMU. Manuscript drafting: IC, IA, UE, CMU. Critical revision: IC, IA, UE, CMU. Supervision: IA, UE, CMU. All authors have discussed and approved the final manuscript. UCM and UE are Joint Senior Authors.

